# NUTRITIONAL KNOWLEDGE AND ASSOCIATED FACTORS AMONG PREGNANT WOMEN IN GHANA: A CROSS-SECTIONAL STUDY

**DOI:** 10.64898/2026.04.13.26350744

**Authors:** Mary Nkansah, Promise Kwame Salu, Linda Afriyie Gyimah

## Abstract

**Background:** Adequate maternal nutritional knowledge is essential for healthy pregnancy outcomes, yet many pregnant women lack good nutritional knowledge. This study assessed nutritional knowledge and associated factors among pregnant women in the Krowor Municipality of Ghana.

**Methods:** A facility-based cross-sectional study was conducted among pregnant women attending antenatal clinics in two public health facilities. Structured questionnaires were used to collect data on sociodemographic characteristics and nutritional knowledge. Data were analysed using descriptive statistics and chi-square tests at a 5% significance level.

**Results:** Most respondents demonstrated moderate nutritional knowledge (mean score =11.24 ± 2.48), with 45% classified as having moderate knowledge. Income level (p = 0.00), education (p = 0.007), gestational age (p = 0.042), employment status (p = 0.007), and religion (p = 0.005) were significantly associated with nutritional knowledge.

**Conclusion:** The study highlights notable gaps in nutritional knowledge among pregnant women in Krowor Municipality. Socioeconomic and obstetric factors strongly influenced nutritional knowledge. Strengthening antenatal nutrition counselling and improving socioeconomic support may help improve the nutritional knowledge of pregnant women.

## Introduction

Adequate nutrition during pregnancy is a critical determinant of both maternal and fetal health. Insufficient intake of key nutrients such as iron, folate, and other micronutrients has been linked to adverse outcomes, including maternal anaemia, low birth weight, preterm birth, and impaired neonatal development [1,2]. Nutritional knowledge plays a vital role in shaping dietary behaviours during pregnancy, influencing food choices, adherence to dietary recommendations, and overall nutritional status [3,4]. Factors such as awareness of nutritious foods, food taboos, women’s empowerment, and pregnancy-related cravings further affect maternal dietary practices and nutritional outcomes [1]. Limited nutrition-related knowledge has been identified as a key contributor to the high burden of undernutrition and micronutrient deficiencies among women, as it directly influences attitudes and eating behaviours [3]. Therefore, adequate nutritional knowledge is essential for promoting healthy dietary practices and improving pregnancy outcomes.

Despite its importance, evidence from low-resource settings indicates that many pregnant women lack sufficient understanding of nutritional requirements during pregnancy, with notable gaps in knowledge of appropriate dietary practices [5,6]. Several socio-demographic and obstetric factors, including educational attainment, socioeconomic status, access to nutrition information, and frequency of antenatal care (ANC) visits, have been shown to influence nutritional knowledge [5,7]. These factors interact in complex ways to shape women’s ability to access, interpret, and apply nutrition-related information.

However, empirical evidence on nutritional knowledge and its associated socio-demographic and obstetric determinants remains limited in the Ghanaian context. This study therefore assessed nutritional knowledge and its associated factors among pregnant women in Ghana. Understanding these relationships is important for identifying knowledge gaps and informing the development of targeted nutrition education interventions aimed at improving maternal nutritional literacy and pregnancy outcomes.

## Materials and methods

### Study design and setting

This study adopted a descriptive cross-sectional design. The research was conducted at two key public health facilities: LEKMA Polyclinic and Clean Town Health Centre which together serve the majority of pregnant women in the municipality. Krowor is a largely urban district with an estimated population of 143,012, half of whom are female, and its rapidly changing urban environment makes it a particularly important setting for examining maternal nutrition behaviours.

### Participants

The study participants consisted of pregnant women aged 20 to 45 years who were attending antenatal clinics at the two facilities. To ensure that participants adequately represented the local maternity population, women were included if they had resided in the municipality for at least one year, were registered for ANC at either facility, and intended to deliver at the same facility. Women attending the facilities for non-ANC purposes, as well as those diagnosed with chronic illnesses that could significantly alter dietary behaviours including HIV, type II diabetes, confirmed malaria, or cardiovascular disease were excluded. Women requiring specialist obstetric care or with a history of recurrent miscarriages were also excluded to avoid confounding influences on dietary practices. The systematic random sampling was employed, to select 266 participants out of 757 women across the two facilities.

### Study variables

Nutritional knowledge refers to an individual’s understanding and awareness of food, nutrients, and healthy eating practices, as well as how these influence overall health and well-being. Socio-demographic variables include age, level of education, employment status, income level, marital status, parity, number of ANC visits, religion, ethnicity, nutritional education, gestational age and source of food.

### Measurements

Data collection was undertaken using a structured and pretested questionnaire administered through face-to-face interviews. The questionnaire captured socio-demographic and obstetric information, nutritional knowledge, and meal skipping. Nutritional knowledge was assessed using 14 yes/no items adapted from Lim et al. [8]. Prior to data collection, the questionnaire was pretested with 13 pregnant women at Korle-Bu Polyclinic to ensure clarity and cultural appropriateness. Necessary revisions were made based on feedback. Data collection took place in private spaces within the ANC from January to March 2023. For participants who could not read or write, the questions were read aloud and responses recorded by trained research assistants.

### Bias

Measures were taken to reduce possible sources of bias in this study. Selection bias was addressed by using the systematic random approach alongside well-defined inclusion criteria. Information bias was minimised through the use of a structured, pre-tested questionnaire to promote consistency in data collection. In addition, social desirability bias was limited by assuring participants of anonymity and confidentiality, and by ensuring that participation was entirely voluntary.

### Sample size

The sample size for this study was determined using the Yamane (1967) formula [10], which provides a simplified approach for calculating sample size for a finite population. With a population of 757 and a margin of error of 0.05, a minimum sample size of approximately 262 respondents was obtained. This was increased to 266 to account for potential non-response.

### Data analysis

Collected data were entered into Microsoft Excel, cleaned, and exported into SPSS version 22 for analysis. Descriptive statistics such as frequencies and percentages were used to summarise participant characteristics and nutritional knowledge. Nutritional knowledge scores were computed and categorised into good (80–100%), moderate (50–79%), and poor (<50%) based on Bloom’s cut-off criteria [9]. The association between socio-demographic factors and nutritional knowledge was examined using chi-square tests, with significance set at p < 0.05. Missing data were handled using mean imputation for quantitative variables and mode imputation for categorical variables.

### Patient and public involvement

Patients were not involved in the design or conduct of this study.

## Results

### Characteristics of the respondents

Table 1 shows information on socio-demographic background of respondents. A total of 266 pregnant women participated in the study. Most participants were between the ages of 20-30 (n = 180, 68 %). Majority of the pregnant women had parity ranging from 0-2 (n = 193, 73%) with a mean of 2.33 (± 0.86). Most participants recorded ANC visits ranging from 1-3 (n= 121, 46%). Majority of the respondents were married (n = 169, 64%). Additionally, most of them had completed basic education (n =101, 38%). A greater proportion were Christians (n = 198, 75%) and came from the Akan ethnic group (n= 96, 36%). Most participants had income levels below GHC 500.00 (n = 73, 45%) and were employed at the time of data collection (n = 191, 72%). A greater proportion had been exposed to nutrition education prior to the study (n = 231, 87%) and were in the second trimester of pregnancy (n = 109, 41%).

**Table 1:**
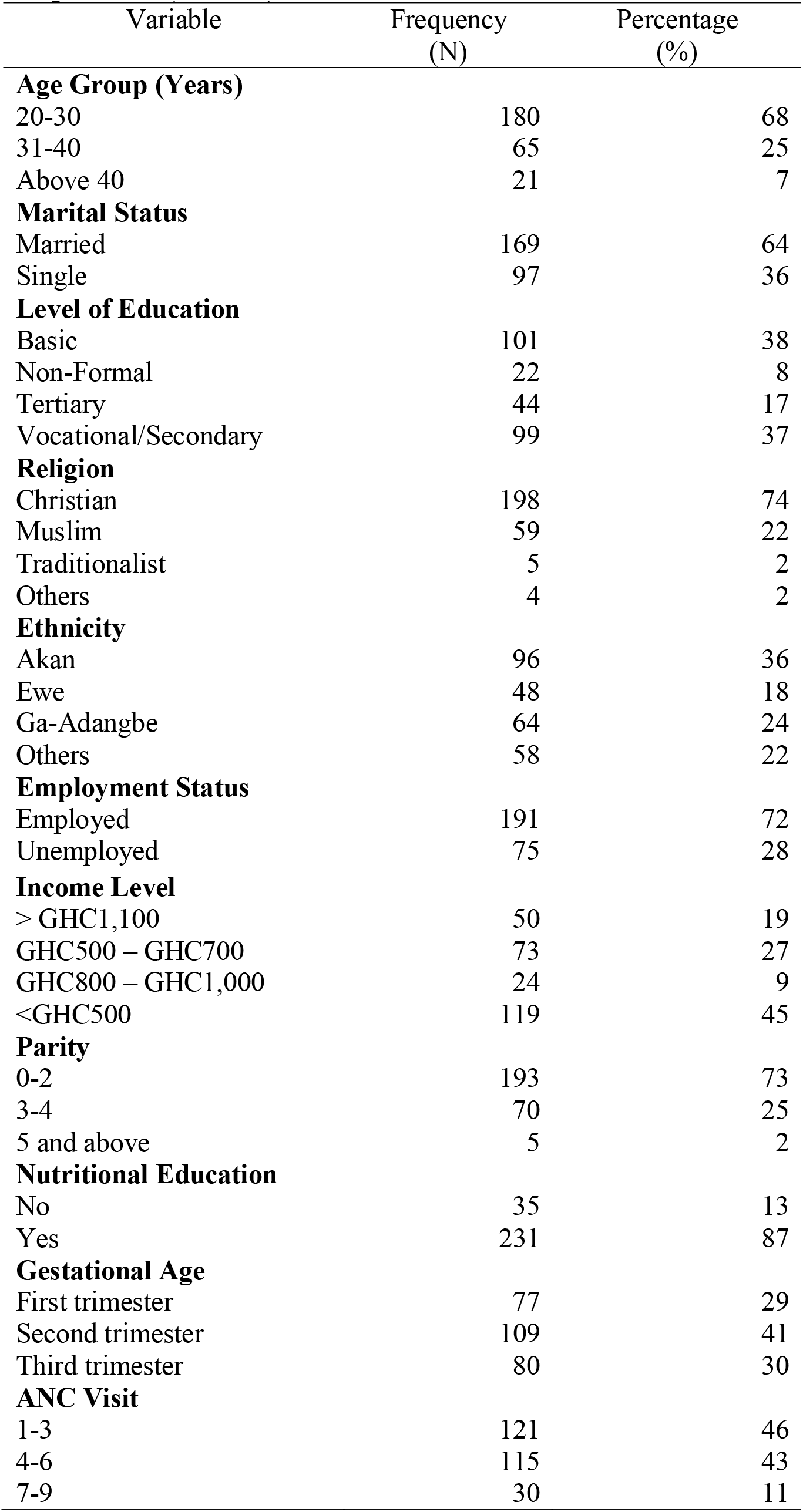
Socio-demographic and obstetric characteristics of respondents (N = 266)

| Variable | Frequency<br>(N) | Percentage<br>(%) |
| --- | --- | --- |
| **Age Group (Years)** |  |  |
| 20-30 | 180 | 68 |
| 31-40 | 65 | 25 |
| Above 40 | 21 | 7 |
| **Marital Status** |  |  |
| Married | 169 | 64 |
| Single | 97 | 36 |
| **Level of Education** |  |  |
| Basic | 101 | 38 |
| Non-Formal | 22 | 8 |
| Tertiary | 44 | 17 |
| Vocational/Secondary | 99 | 37 |
| **Religion** |  |  |
| Christian | 198 | 74 |
| Muslim | 59 | 22 |
| Traditionalist | 5 | 2 |
| Others | 4 | 2 |
| **Ethnicity** |  |  |
| Akan | 96 | 36 |
| Ewe | 48 | 18 |
| Ga-Adangbe | 64 | 24 |
| Others | 58 | 22 |
| **Employment Status** |  |  |
| Employed | 191 | 72 |
| Unemployed | 75 | 28 |
| **Income Level** |  |  |
| > GHC1,100 | 50 | 19 |
| GHC500 – GHC700 | 73 | 27 |
| GHC800 – GHC1,000 | 24 | 9 |
| <GHC500 | 119 | 45 |
| **Parity** |  |  |
| 0-2 | 193 | 73 |
| 3-4 | 70 | 25 |
| 5 and above | 5 | 2 |
| **Nutritional Education** |  |  |
| No | 35 | 13 |
| Yes | 231 | 87 |
| **Gestational Age** |  |  |
| First trimester | 77 | 29 |
| Second trimester | 109 | 41 |
| Third trimester | 80 | 30 |
| **ANC Visit** |  |  |
| 1-3 | 121 | 46 |
| 4-6 | 115 | 43 |
| 7-9 | 30 | 11 |

Over two-thirds were into trading (39%). Figure 1 provides a breakdown of specific occupations of respondents.

**Figure 1:**
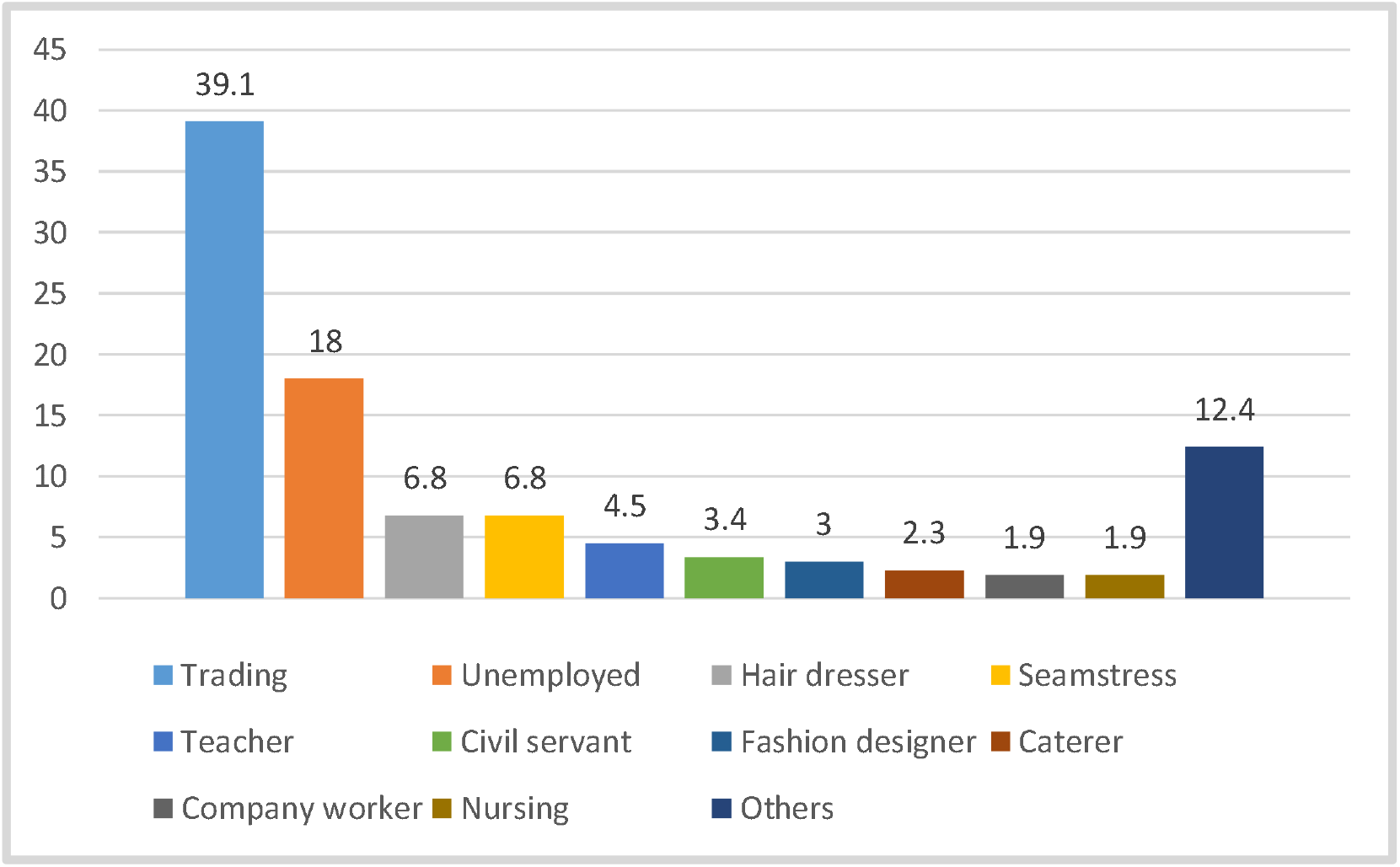
Occupational Background of Respondents.

### Level of nutritional knowledge

Figure 2 depicts the level of nutritional knowledge among respondents. The mean knowledge score was 11.24 (±2.48) on a 20-point scale. Most of the participants had moderate level of nutritional knowledge (45%).

**Figure 2:**
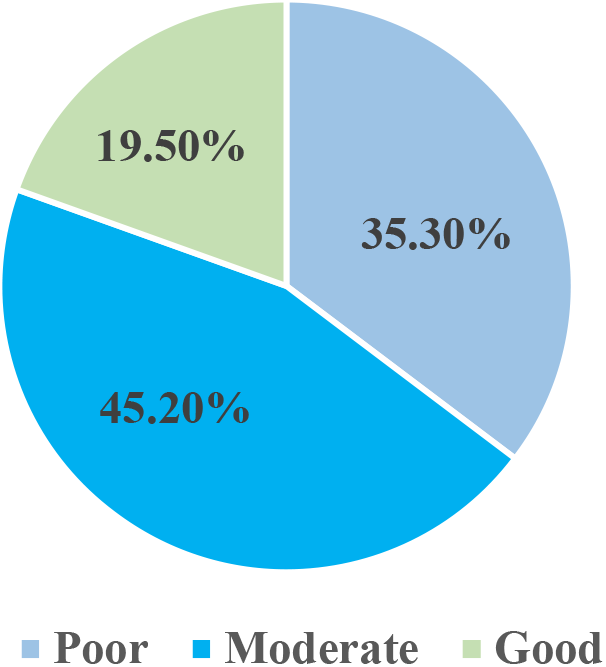
Nutritional knowledge of pregnant women.

### Factors associated with nutritional knowledge

Table 2 shows the factors associated with nutritional knowledge. Income level (p = 0.00), level of education (p = 0.007), gestational age (p= 0.042), employment status (p = 0.007) and religion (p = 0.005) presented a significant association with nutritional knowledge.

**Table 2:**
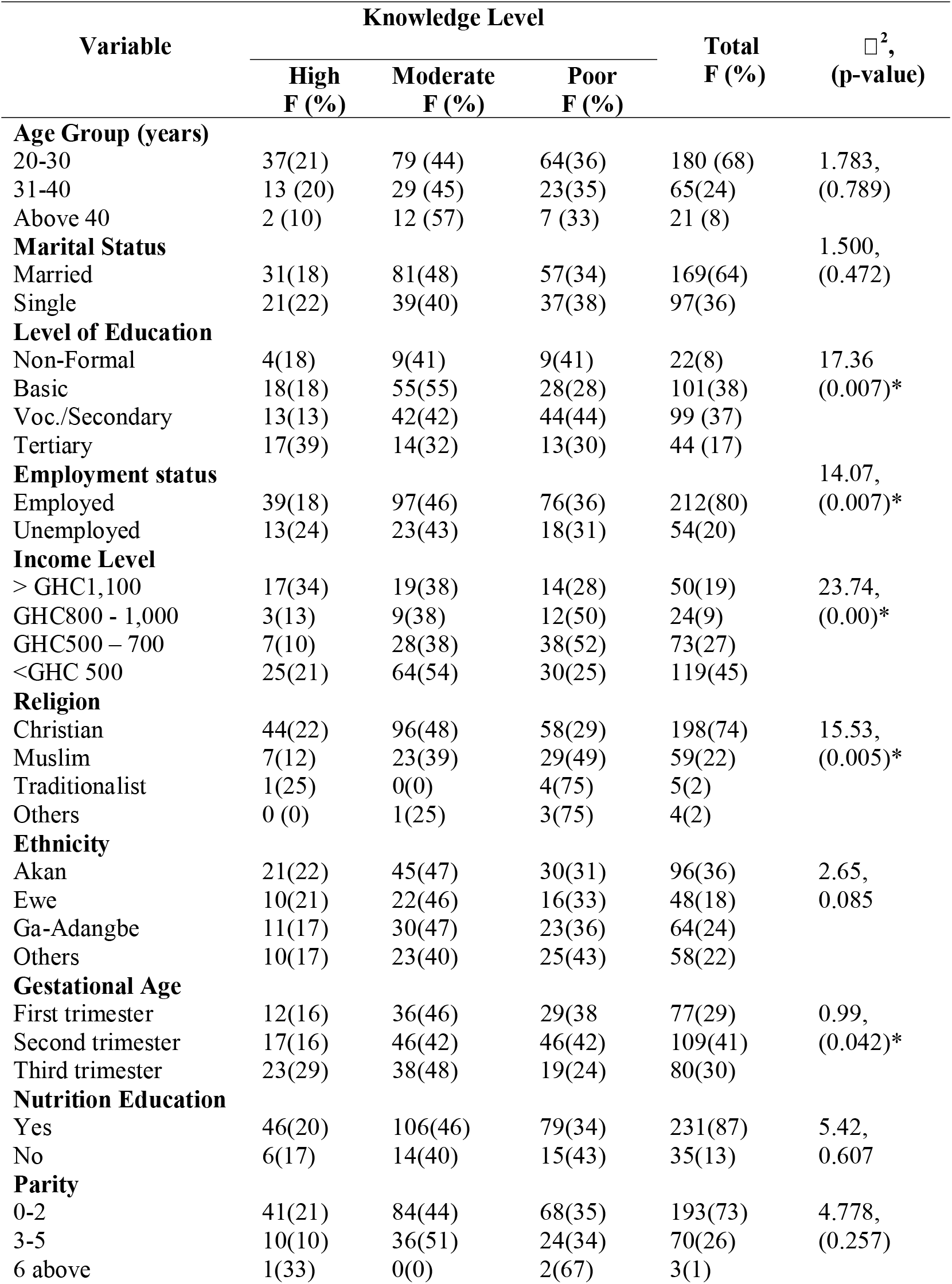
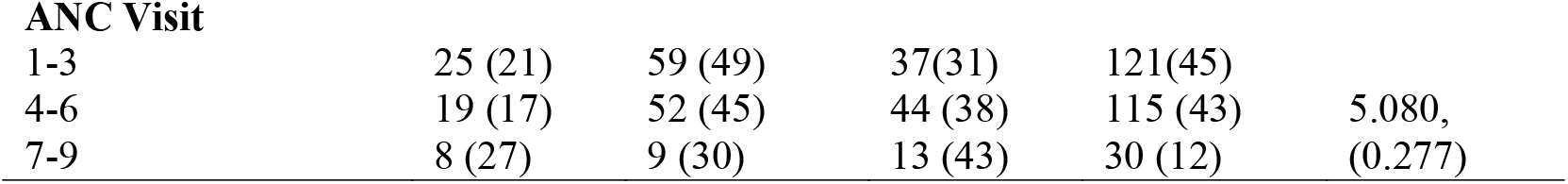
Chi-square analysis of factors associated with nutritional knowledge.

| Variable | Knowledge Level | | | Total<br>F (%) | $\chi^2$,<br>(p-value) |
| --- | --- | --- | --- | --- | --- |
|  | High<br>F (%) | Moderate<br>F (%) | Poor<br>F (%) |  |  |
| **Age Group (years)** |  |  |  |  |  |
| 20-30 | 37(21) | 79 (44) | 64(36) | 180 (68) | 1.783, |
| 31-40 | 13 (20) | 29 (45) | 23(35) | 65(24) | (0.789) |
| Above 40 | 2 (10) | 12 (57) | 7 (33) | 21 (8) |  |
| **Marital Status** |  |  |  |  | 1.500, |
| Married | 31(18) | 81(48) | 57(34) | 169(64) | (0.472) |
| Single | 21(22) | 39(40) | 37(38) | 97(36) |  |
| **Level of Education** |  |  |  |  |  |
| Non-Formal | 4(18) | 9(41) | 9(41) | 22(8) | 17.36 |
| Basic | 18(18) | 55(55) | 28(28) | 101(38) | (0.007)* |
| Voc./Secondary | 13(13) | 42(42) | 44(44) | 99 (37) |  |
| Tertiary | 17(39) | 14(32) | 13(30) | 44 (17) |  |
| **Employment status** |  |  |  |  | 14.07, |
| Employed | 39(18) | 97(46) | 76(36) | 212(80) | (0.007)* |
| Unemployed | 13(24) | 23(43) | 18(31) | 54(20) |  |
| **Income Level** |  |  |  |  |  |
| > GHC1,100 | 17(34) | 19(38) | 14(28) | 50(19) | 23.74, |
| GHC800 - 1,000 | 3(13) | 9(38) | 12(50) | 24(9) | (0.00)* |
| GHC500 – 700 | 7(10) | 28(38) | 38(52) | 73(27) |  |
| <GHC 500 | 25(21) | 64(54) | 30(25) | 119(45) |  |
| **Religion** |  |  |  |  |  |
| Christian | 44(22) | 96(48) | 58(29) | 198(74) | 15.53, |
| Muslim | 7(12) | 23(39) | 29(49) | 59(22) | (0.005)* |
| Traditionalist | 1(25) | 0(0) | 4(75) | 5(2) |  |
| Others | 0 (0) | 1(25) | 3(75) | 4(2) |  |
| **Ethnicity** |  |  |  |  |  |
| Akan | 21(22) | 45(47) | 30(31) | 96(36) | 2.65, |
| Ewe | 10(21) | 22(46) | 16(33) | 48(18) | 0.085 |
| Ga-Adangbe | 11(17) | 30(47) | 23(36) | 64(24) |  |
| Others | 10(17) | 23(40) | 25(43) | 58(22) |  |
| **Gestational Age** |  |  |  |  |  |
| First trimester | 12(16) | 36(46) | 29(38) | 77(29) | 0.99, |
| Second trimester | 17(16) | 46(42) | 46(42) | 109(41) | (0.042)* |
| Third trimester | 23(29) | 38(48) | 19(24) | 80(30) |  |
| **Nutrition Education** |  |  |  |  |  |
| Yes | 46(20) | 106(46) | 79(34) | 231(87) | 5.42, |
| No | 6(17) | 14(40) | 15(43) | 35(13) | 0.607 |
| **Parity** |  |  |  |  |  |
| 0-2 | 41(21) | 84(44) | 68(35) | 193(73) | 4.778, |
| 3-5 | 10(10) | 36(51) | 24(34) | 70(26) | (0.257) |
| 6 above | 1(33) | 0(0) | 2(67) | 3(1) |  |
| **ANC Visit** |  |  |  |  |  |
| 1-3 | 25 (21) | 59 (49) | 37(31) | 121(45) |  |
| 4-6 | 19 (17) | 52 (45) | 44 (38) | 115 (43) | 5.080, |
| 7-9 | 8 (27) | 9 (30) | 13 (43) | 30 (12) | (0.277) |

## Discussion

This study found that most pregnant women possessed moderate nutritional knowledge. Levels of income and education, gestational age, employment status and religion were found to be significantly associated with nutritional knowledge.

The moderate nutritional knowledge demonstrated by participants suggests that while many pregnant women are aware of basic nutrition concepts, important knowledge gaps remain. Sub-optimal nutritional knowledge among pregnant women may result from limited access to quality antenatal nutrition education, particularly in settings where counselling during antenatal care is inconsistent [6,11]. In addition, inadequate exposure to reliable health information through mass media or community-based programmes may restrict awareness of appropriate dietary practices during pregnancy [12]. Socioeconomic constraints, including low household income and food insecurity, can also limit women’s ability to engage with or prioritise nutrition-related information, thereby indirectly affecting knowledge acquisition [13].

Studies across Africa indicate that nutritional knowledge among pregnant women is often suboptimal, with many women demonstrating moderate levels of knowledge. In Ghana, a study reported moderate nutrition knowledge among pregnant women [14]. Similarly, another study revealed important knowledge gaps, with only 16.9% of respondents aware of the importance of folic acid supplementation [13]. Comparable findings have been reported in Ethiopia, where many pregnant women were found to have inadequate nutritional knowledge [7,5]. However, this finding contradicts that of Appiah et al., which reported higher nutritional knowledge among adolescent pregnant women in Ghana [15]. The difference in outcome might result from differences in study characteristics. While the study by Appiah et al. [15] involved 423 participants, the present study involved 266 participants. Furthermore, while the present study involved older adults, the study by Appiah et al. [15] comprised of adolescents.

The associations between levels of income and education, gestational age, employment status, religion and nutritional knowledge reflect the complex social and economic realities that shape women’s ability to access, understand, and apply nutrition information during pregnancy. The association between income level and nutritional knowledge emphasises the significance of socio-economic factors to the availability of nutrition education and awareness. The finding is supported by previous studies which reported that higher income enhances access to education and health information, thereby facilitating better understanding of appropriate dietary practices [11,12]. Similarly, the relationship between education level and nutritional knowledge highlights the crucial role that education plays in shaping individuals’ understanding of dietary principles and practices. This aligns with previous studies which reports that formal education and access to nutritional information significantly increase the odds of being knowledgeable [15–17].

Furthermore, the observed association between gestational age and nutrition knowledge is not unexpected because as the pregnancy advances, the women may have actively sought out information and resources regarding nutrition in order to support the developing foetus and address their own evolving nutritional requirements. Gestational age may influence nutritional knowledge, as women in later stages of pregnancy are more likely to have attended multiple antenatal care (ANC) visits and received repeated nutrition counselling. Evidence shows that ANC provides structured opportunities for nutrition education and behaviour change, thereby improving women’s knowledge and dietary practices over time [18]. In addition, studies indicate that pregnant women often receive and recall dietary information progressively across antenatal visits, suggesting that continued exposure enhances knowledge acquisition as pregnancy advances [19].

The association between employment status and nutritional knowledge might suggest that employed women have better access to information which can translate into higher nutrition knowledge scores. Employment often correlates with higher income and education, both of which facilitate exposure to health information through media, workplace interactions, and more regular use of health services [16]. However, this association should be interpreted with caution, as it may be confounded by factors such as educational attainment and income. Evidence shows that maternal education and household wealth are significantly associated with improved nutritional knowledge, as they enhance health literacy and access to information [1,20]. Women with higher education are more likely to be employed and to possess better understanding of health information. In addition, improved access to antenatal care services and health information has been shown to significantly influence nutritional knowledge, suggesting that employed women may benefit from greater exposure to counselling and health services [20]. Furthermore, occupation itself has been identified as a determinant of nutritional knowledge, with employed or professional women demonstrating higher knowledge levels compared to unemployed women [21]. Reverse causality is also possible, whereby women with better nutritional knowledge may be more likely to seek or maintain employment, although causal relationships remain difficult to establish in cross-sectional studies [20]. The finding aligns with previous work that links higher socioeconomic position to improved maternal nutrition knowledge and practices [1,22].

### Strengths and limitations of the study

The study is strengthened by its use of systematic sampling, adequate sample size, pre-tested data collection tools, and rigorous bias control measures. It also incorporates comprehensive variables and appropriate statistical analyses, while providing context-specific evidence relevant to maternal nutrition in Ghana. Ethical standards were maintained, and the findings have clear public health relevance. The study however has some limitations. First, the study was conducted in selected public health facilities, which may limit the generalisability of the findings to pregnant women who do not attend antenatal care or those in private or rural settings. Furthermore, although a structured questionnaire was used, it may not have fully captured the depth of participants’ nutritional knowledge and underlying beliefs. Some potential confounding factors, such as household food security, cultural influences, and partner support, were also not extensively assessed. However, these limitations are unlikely to have significantly affected the overall validity of the findings, as appropriate methodological measures, including systematic sampling and the use of a pre-tested instrument, were employed to enhance the reliability and credibility of the results.

## Conclusion

This study assessed the level of nutritional knowledge and associated factors among pregnant women. The findings revealed that a considerable proportion of respondents had moderate nutritional knowledge. This result suggests the presence of important gaps in maternal nutrition awareness. Strengthening nutrition education during antenatal care is essential to enhance maternal and fetal health outcomes. Future research should explore context-specific interventions to improve nutrition-related behaviours among pregnant women.

## Data Availability

All data produced in the present study are available upon reasonable request to the authors.

## Author’s Contributions

Conceptualisation: MN, LAG. Data collection: MN, PKS. Result analysis and presentation: MN, LAG, PKS. Writing of original draft PKS. All authors contributed to the review and editing of manuscript and approved the final draft.

## Ethical approval and consent to participate

Ethical approval was obtained from the Ghana Health Service Ethics Review Committee (GHS-ERC: 050/09/22). Participants signed informed consent forms to indicate their voluntary participation in the study.

## Consent for publication

Not applicable

## Availability of data

The datasets used and/or analysed during the current study are available from the corresponding author on reasonable request.

## Acknowledgement

We acknowledge the Greater Accra Regional Health Directorate, the LEKMA Health Directorate and the hospital directors of LEKMA Polyclinic and Clean Town Health Centre for granting us access to their facilities for the study.

## References

1. Gebremichael MA, Assefa N, Bekele T, et al. Prevalence and predictors of knowledge and attitude on optimal nutrition among pregnant women. Int J Womens Health 2023;15:893–906. doi:10.2147/IJWH.S415615

2. UNICEF. Maternal nutrition: improving maternal and newborn health and survival. New York: UNICEF; 2022. Available from: https://www.unicef.org/health/maternal-and-newborn-health

3. Weerasekara PC, Withanachchi CR, Ginigaddara GAS, Ploeger A. Food and nutrition-related knowledge among women. Int J Environ Res Public Health 2020;17(11):3985. doi:10.3390/ijerph17113985

4. Katenga-Kaunda LZ, Kamudoni PR, Holmboe-Ottesen G, et al. Enhancing nutrition knowledge among pregnant women. BMC Pregnancy Childbirth 2021;21:644. doi:10.1186/s12884-021-04117-5

5. Demisew M, Gemede HF, Ayele K. Knowledge and attitude towards nutrition among pregnant women. J Nutr Sci 2024;13:e23. doi:10.1017/jns.2024.19

6. Zerfu TA, Biadgilign S. Limited knowledge and poor dietary diversity practices. BMC Nutr 2018;4:1–9. doi:10.1186/s40795-018-0251-x

7. Gezimu W, Bekele F, Habte G. Nutrition knowledge and practice among pregnant mothers. SAGE Open Med 2022;10:20503121221085843. doi:10.1177/20503121221085843

8. Lim ZX, Wong JL, Lim PY, Soon LK. Knowledge of nutrition during pregnancy. Int J Public Health Clin Sci 2018;5(1):117–128.

9. Bloom BS. Taxonomy of educational objectives. New York: Longmans; 1956.

10. Yamane T. Statistics: an introductory analysis. 2nd ed. New York: Harper & Row; 1967.

11. Girard AW, Olude O. Nutrition education and counselling during pregnancy. Paediatr Perinat Epidemiol 2012;26(Suppl 1):191–204. doi:10.1111/j.1365-3016.2012.01278.x

12. Kavle JA. Strengthening maternal nutrition counselling. Public Health Nutr 2023;26(2):363–380. doi:10.1017/S1368980022002129

13. Agyei EA, Afrifa SK, Munkaila A, et al. Income level and dietary diversity among pregnant women. J Nutr Metab 2021;2021:5581445. doi:10.1155/2021/5581445

14. Adjei-Banuah NY, Aduah VA, Ziblim SD, et al. Nutrition knowledge among pregnant women in Ghana. Nutr Metab Insights 2021;14:1–9. doi:10.1177/11786388211039427

15. Appiah PK, Naa Korklu AR, Bonchel DA, et al. Nutritional knowledge among pregnant adolescents. J Nutr Metab 2021;2021:8835704. doi:10.1155/2021/8835704

16. Tesfaye T, Dadi AF, et al. Nutritional knowledge among pregnant adolescents. Sci Rep 2024;14:57428. doi:10.1038/s41598-024-57428-w

17. Yeneabat T, Adugna HH, Asmamaw T, et al. Maternal dietary diversity and micronutrient adequacy. BMC Pregnancy Childbirth 2019;19:173. doi:10.1186/s12884-019-2299-2

18. Bayked EM, Yimer EM, Gelaw T, et al. Dietary knowledge and associated factors among pregnant mothers. Front Public Health 2024;12:1393764. doi:10.3389/fpubh.2024.1393764

19. Heri R, Malqvist M, Yahya-Malima KI, et al. Dietary diversity and associated factors among women attending antenatal clinics in Tanzania. BMC Nutr 2024;10:16. doi:10.1186/s40795-024-00825-1

20. Bryant J, Hure A, et al. Receipt of information about diet by pregnant women: a cross-sectional study. Women Birth 2019;32(6):e501–e507. doi:10.1016/j.wombi.2018.12.005

21. Marian M, Pérez RL, McClain AC, et al. Nutritional knowledge and practices of low-income women during pregnancy. J Health Popul Nutr 2025;44:33. doi:10.1186/s41043-025-00776-8

22. Oladebo T, Bobholz F, Folivi K, et al. A scoping review on nutrition knowledge and nutrition literacy among pregnant women and the prevalence of pregnancy complications and adverse pregnancy outcomes. Nutrients 2025;17(21):3488. doi:10.3390/nu17213488

